# Opposite Directions: A Decade of Contrasting HIV and HCV Dynamics Among Injecting Drug Users in Mozambique

**DOI:** 10.64898/2026.03.09.26347977

**Authors:** Áuria Ribeiro Banze, Rachid Muleia, Lúcia Muioche, Samuel Nuvunga, Gércio Cuamba, Manuel Condula, Stélio Craveirinha, Diogo Chavana, Ana Mutola Jemuce, Valdo Mega, Daniel Chilaúle, Maria Helena Simbine, Carlos Botão, Nália Ismael, Cynthia Semá Baltazar

## Abstract

People who inject drugs (PWID) experience a high burden of HIV and hepatitis C virus (HCV) infection due to unsafe injection practices and limited access to harm-reduction services. In Mozambique, data on PWID remain limited. This study analyzed two rounds of biobehavioral surveys conducted in 2014 and 2023 in Maputo and Nampula to assess trends in HIV and HCV prevalence and to identify associated behavioral and structural factors. We compared unweighted prevalence estimates using descriptive analysis and applied multivariable logistic regression to examine independent associations with each infection and interaction effects with survey year. HIV prevalence declined across most demographic and behavioral groups. Among PWID aged ≥25 years, prevalence decreased from 55.7% to 26.3%, and among men from 45.7% to 16.7% (both p < 0.001). Reductions were also observed among daily injectors (58.0% to 21.3%) and individuals reporting syringe sharing (75.0% to 21.8%). In Maputo, HIV prevalence declined from 56.6% to 28.0%, while the decrease in Nampula was not statistically significant. Age and female sex were strong predictors of HIV infection in the earlier survey, although the association with age weakened in 2023. HCV prevalence showed divergent trends. In Maputo, prevalence decreased from 49.3% to 18.7% (p < 0.001), whereas in Nampula it increased from 11.7% to 48.1% (p < 0.001). PWID aged 16–24 years experienced a fivefold increase in HCV prevalence. Interaction analysis demonstrated a significant rise in Nampula in 2023 (AOR 14.6; p < 0.001). Lower injection frequency and not sharing needles were protective factors for both HIV and HCV. These findings indicate a substantial reduction in HIV prevalence among PWID in Mozambique over the past decade, alongside an increase in HCV prevalence in specific geographic and age groups. The contrasting trends highlight the need for differentiated harm-reduction strategies, expansion of HCV prevention and treatment services, and tailored interventions for subgroups at elevated risk.

## Introduction

Injecting drug use is one of the fastest-changing and least-monitored drivers of HIV and viral hepatitis in sub-Saharan Africa, yet it remains largely invisible in national surveillance systems. This epidemiological blind spot persists despite global commitments to end AIDS and eliminate viral hepatitis as public health threats by 2030. In 2024, an estimated 40.8 million people living with HIV (PLHIV), nearly two-thirds of whom reside in Africa [1].

Hepatitis C virus (HCV) continues to contribute substantially to global morbidity and mortality (5,6). His elimination, although achievable, would prevent more than 1.2 million deaths annually but require significant economic and health-system investment. Their elimination, although achievable, would prevent more than 1.2 million deaths annually but require significant economic and health-system investment. Injecting drug use plays a central role in sustaining these epidemics (4). Co-infection of HIV and HCV remains a major clinical and programmatic challenge, with 6% of PLHIV affected due to shared sexual, blood-borne and injection-related transmission routes [2]. People who inject drugs (PWID) sit at the intersection of these overlapping epidemics [3]. Globally, an estimated 14 million PWID, of whom 1.7 million are living with HIV and 49.3% have viremic HCV infection [4]. Injecting drug use is responsible for roughly 8% of all new HIV infections globally [5] and between 23% and 39% of new HCV infections [6]. Yet, PWID across sub-Saharan Africa remains underserved by harm reduction and clinical services, leaving major gaps in prevention, testing and treatment.

Mozambique is considered a high HIV burden country, with an estimated prevalence of 12.5% among individuals aged 15-49 years old, reported by the Population-based HIV Impact Assessment (PHIA) survey conducted in 2021 [7]. The Mozambique National HIV Strategic Plan (PEN) recognizes PWID as a priority group for targeted HIV/AIDS interventions [8]. However, data on HIV and HCV among PWID remain extremely limited. The first Bio-Behavioral Surveys (BBS) conducted in 2013–2014 revealed alarmingly high HIV prevalence, over 50% in Maputo and nearly 20% in Nampula/Nacala [3]. A second BBS conducted in 2023 offers a rare opportunity, very few countries in the region possess two comparable rounds of biological and behavioral data among PWID spanning an entire decade.

This unique dataset enables the first longitudinal assessment of HIV, HBV and HCV burden among PWID in Mozambique. Yet, critical questions remain regarding how these epidemics are evolving, which subgroups are most affected, and what behavioral and structural factors are driving risk. Understanding these dynamics is essential at a moment when global elimination strategies depend on reaching populations historically left behind. Therefore, this study aims to evaluate changes in the prevalence of HIV and HCV, as well as associated risk behaviors over time, thereby generating evidence to inform targeted prevention and harm reduction strategies for this key population.

## Materials and methods

### Study design

Data analysis from two cross-sectional survey rounds among PWID conducted in Mozambique was performed: the first round in 2013-2014 [3] and the second in 2023-2024[9]. The first BBS was carried out in two primary urban areas Maputo and Nampula while the subsequent 2023 BBS was extended to include three additional cities, Beira, Tete and Quelimane. Recruitment for the second survey was conducted between 01/07/2023 and 31/03/2024. Both surveys collected information on socio-demographic characteristics, sexual and drug use behaviors, injection-related practices (including needle and syringe sharing), and access to health services among PWID.

### Study population and sample size

The 2014 BBS allowed individuals aged 18 years and above while the and 2023 BBS, PWID were eligible to participate if they were at least 16 years old, had injected drugs without medical prescription in the past 12 months, resided, worked, or socialized in the survey area during the same period, presented a valid referral coupon, and were able to provide informed consent. These criteria ensured the inclusion of active PWID who were socially connected within the study areas. For comparability purposes, this analysis considered only a subsample of participants aged 18 years and above.

The first round of BBS, 353 and 139 were recruited for Maputo and Beira, respectively During the second round, 520 were recruited in Maputo, 517 in Beira, 524 in Tete, 531 in Quelimane and 532 in Nampula. For the current data analysis set, a total of 492 PWID from the 2014 BBS, and 2624 PWID from the 2023 BBS with valid HIV tests were used. In both surveys, study participants were recruited using RDS, a methodology designed to effectively sample populations that are traditionally challenging to access. More details about the survey methodology are described elsewhere [10].

### Data collection and measures

Socio-behavioral data were collected using a standardized questionnaire that captured information on socio-demographic characteristics, drug use history and injection practices, sharing of injection equipment, sexual behaviors (including sexual history, number of partners, and condom use), alcohol and other substance use, access to and utilization of harm reduction and healthcare services, as well as knowledge of HIV, hepatitis, and related health conditions.

HIV testing followed the national sequential algorithm, with Alere Determine® HIV-1/2 as the screening test and Uni-Gold™ HIV for confirmation. Results were indeterminate if the confirmatory test was non-reactive. HCV infection was assessed using the SD Bioline HCV rapid antibody test, with all reactive results considered positive for the purpose of prevalence estimation All participants received pre- and post-test counselling with personalized messages according to their test results and risk profiles. Those with positive results for HIV or HCV were referred to appropriate care services. Referral forms included anti-HCV results to facilitate clinical linkage and confirmatory assessment at designated health facilities.

### Ethics approval and consent to participate

The 2014 and 2023 Bio-Behavioural Surveys received ethical approval from the National Bioethics Committee for Health (CNBS) in Mozambique (references 877/CNBS/22 and 097/CIBS-INS/2022). Approval was also granted by the Institutional Ethics Committee of the National Institute of Health (CIE-INS). Written informed consent was obtained from all eligible participants prior to data collection. For each survey, participants could choose to provide consent for the questionnaire only or for both the questionnaire and rapid diagnostic testing.

This secondary analysis was authorized by the INS Data Management Unit and conducted in accordance with national ethical regulations and internationally recognized standards.

### Statistical Analysis

In this study we conducted both descriptive and multivariate analysis. The descriptive analysis served to describe the study sample and to compute the prevalence of the outcome variables with respect to the explanatory variables. Although the data were collected using respondent-driven sampling (RDS), point estimates and confidence intervals were calculated without weighting because the sample was aggregated across multiple cities, and RDS estimators are valid only within individual recruitment networks. For comparison of prevalences of HIV between the two surveys, we used a Chi-square test for equality of proportion. Multivariate analyses were conducted through logistic regression to understand how selected explanatory variables are related to the three outcome variables, HIV and HCV status. Additionally, we used to evaluate how the risk of infection of HIV and HCV changed in nine years. To evaluate temporal variation in covariate effects, we included pairwise interactions between survey year and each explanatory variable. Stepwise, variable selection method was applied to remove less important variables and interactions in the multivariate logistic regression. In this analysis, we considered unweighted logistic regression, as many studies suggest that this approach outperforms logistic regression with RDS. All the analysis in this study were conducted using R statistical software version 4.4.1. Statistical significance was evaluated at 5% significance level.

## Results

### Trends in HIV and HCV prevalence by characteristics and risk factors

Table 1 presents the descriptive analysis for BBS implemented in 2014 and 2023 showing the changes in HIV and HCV prevalence according to the selected characteristics.

**Table 1.** Distribution of HIV and HCV prevalence by selected variables for 2014 and 2023 BBS^a^.

| Characteristics | HIV |  |  |  | HCV |  |  |  |
| --- | --- | --- | --- | --- | --- | --- | --- | --- |
|  | 2014 HIV n/N (%) | 2023 HIV n/N (%) | Difference of prevalences % (95% CI)<sup>c</sup> | p-value<sup>b</sup> | 2014 HCV n/N (%) | 2023 HCV n/N (%) | Difference of prevalences % (95% CI)<sup>c</sup> | p-value<sup>b</sup> |
| ***Sociodemographic characteristics*** |  |  |  |  |  |  |  |  |
| Age group: 18-24 | 5/81(6.2) | 42/322(13.0) | 6.9 ( -0.3; 14) | 0.126 | 6/82(7.3) | 126/326(38.7) | 31.3 (22.8; 39.8) | 0 |
| Age group: ≥25 | 190/341(55.7) | 156/594(26.3) | -29.5 ( -36; -22.9) | 0 | 154/340(45.3) | 172/597(28.8) | -16.5 (-23.1; -9.8) | 0 |
| Sex: Male | 184/403(45.7) | 125/747(16.7) | -29.2 (-34.8; -23.6) | 0 | 153/406(37.7) | 270/752(35.8) | -1.8 ( -7.8; 4.2) | 0.592 |
| Sex: Female<sup>d</sup> | 11/19(57.9) | 73/169(43.2) | -15.8 (-41.3; 9.8) | 0.257 | 7/16(43.8) | 28/171(16.4) | -27.4 (-55.7; 1) | 0.019 |
| Education: No formal/Primary | 93/179(52.0) | 85/278(30.6) | -21.4 ( -30.9; -11.8) | 0 | 74/172(43.0) | 78/281(27.8) | -15.3 (-24.8; -5.7) | 0.001 |
| Education: Secondary/Higher | 102/242(42.1) | 112/635(17.6) | -24.5 (-31.7; -17.3) | 0 | 86/249(34.5) | 218/639(34.1) | -0.4 ( -7.7; 6.8) | 0.968 |
| Marital status: Never married | 111/247(44.9) | 121/672(18.0) | -26.9 (-34.1; -19.8) | 0 | 93/246(37.8) | 225/678(33.2) | -4.6 (-11.9; 2.7) | 0.219 |
| Marital status: Married/Living in union | 34/86(39.5) | 51/150(34.0) | -5.5 ( -19.3; 8.2) | 0.477 | 25/88(28.4) | 44/150(29.3) | 0.9 (-11.9; 13.7) | 0.997 |
| Marital status: Other | 50/89(56.2) | 26/93(28.0) | -28.2 ( -43.1; -13.4) | 0 | 42/88(47.7) | 28/94(29.8) | -17.9 ( -33; -2.9) | 0.020 |
| City of residence: City Maputo | 171/302(56.6) | 138/493(28.0) | -28.6 (-35.7; -21.5) | 0 | 145/294(49.3) | 93/497(18.7) | -30.6 (-37.5; -23.7) | 0 |
| City of residence: City Nampula/Nacala | 24/120(20.0) | 60/423(14.2) | -5.8 (-14.2; 2.6) | 0.158 | 15/128(11.7) | 205/426(48.1) | 36.4 (28.6; 44.2) | 0 |
| ***Drug use characteristics*** |  |  |  |  |  |  |  |  |
| Age first drug: <18 | 18/49(36.7) | 32/135(23.7) | -13 (-29.7; 3.6) | 0.117 | 15/50(30.0) | 45/137(32.8) | 2.8 (-13.5; 19.2) | 0.848 |
| Age first drug: 18-24 | 86/203(42.4) | 89/476(18.7) | -23.7 (-31.7; -15.7) | 0 | 74/203(36.5) | 179/481(37.2) | 0.8 ( -7.5; 9) | 0.919 |
| Age first drug: ≥25 | 91/170(53.5) | 77/303(25.4) | -28.1 (-37.5; -18.7) | 0 | 71/169(42.0) | 74/303(24.4) | -17.6 (-26.9; -8.3) | 0 |
| Injection frequency: Daily | 145/250 (58.0) | 181 / 851(21.3) | -36.7 (-43.7; -29.8) | 0 | 127/ 245(51.8) | 280/858(32.6) | -19.2 (-26.5; -11.9) | 0 |
| Injection frequency: Weekly/monthly | 50/172 (29.1) | 17/65(26.2) | -2.9 (-16.6; 10.8) | 0.777 | 33/177(18.6) | 18/65(27.7) | 9 (-4.3; 22.4) | 0.176 |
| Access to new syringes: Yes | 171/360(47.5) | 172/812 (21.2) | -26.3 (-32.4; -20.2) | 0 | 136/ 361(37.7) | 279/818(34.1) | -3.6 ( -9.7; 2.6) | 0.265 |
| Access to new syringes: No | 23/61(37.7) | 25/102 (24.5) | -13.2 (-29.3; 2.9) | 0.107 | 24/60(40.0) | 19/103(18.4) | -21.6 (-37.4; -5.8) | 0.005 |
| Shared syringes (past 12 months): Yes | 15/20 (75.0) | 148/696 (21.8) | -53.7 (-75.5; -31.9) | 0 | 15/20(75.0) | 260/669(38.90) | -36.1 ( -58; -14.2) | 0.003 |
| Shared syringes (past 12 months): No | 178/407 (43.7) | 51/246 (20.7) | -23 (-30.3; -15.7) | 0 | 142/391(36.3) | 37/248(14.9) | -21.4 (-28.2; -14.6) | 0 |
| Ever injected with previously used syringe: Yes | 24/45(53.3) | 75/373(20.1) | -33.2 (-49.6; -16.8) | 0 | 15/44(34.1) | 168/375(44.8) | 10.7 ( -5.4; 26.9) | 0.232 |
| Ever injected with previously used syringe: No | 146/320(45.6) | 121/537(22.5) | -23.1 (-29.8; -16.3) | 0 | 123/322(38.2) | 129/542(23.8) | -14.4 ( -21.1; -7.7) | 0 |
| Ever shared any other injection equipment: Yes | 68/142(47.9) | 78/316(24.7) | -23.2 (-33.2; -13.2) | 0 | 54/142(38.0) | 70/319(21.9) | -16.1 (-25.8; -6.4) | 0 |
| Ever shared any other injection equipment: No | 107/237(47.1) | 113/572(19.8) | -25.4 ( -32.8; -18) | 0 | 86/237(36.3) | 215/576(37.3) | 1 (-6.5; 8.6) | 0.842 |
| **Sexual behaviours** |  |  |  |  |  |  |  |  |
| Number of sexual partners (past 12 months): 0-1 | 86/170(50.6) | 83/340(24.4) | -26.2 (-35.4; -16.9) | 0 | 61/165(37.0) | 124/344(38.4) | -0.9 (-10.3; 8.5) | 0.917 |
| Number of sexual partners (past 12 months): 2 | 26/56(46.4) | 25/146(17.1) | -29.3 ( -45; -13.6) | 0 | 27/59(45.8) | 56/146(38.4) | -7.4 (-23.6; 8.7) | 0.412 |
| Number of sexual partners (past 12 months): ≥3 | 83/196(42.3) | 87/414(21.0) | -21.3 (-29.7; -13) | 0 | 72/198(36.4) | 113/416(27.2) | -9.2 (-17.5; -0.9) | 0.026 |
| Unprotected sex (past 12 months): Yes | 66/184 (35.9) | 109/510(21.4) | -14.5 ( -22.7; -6.3) | 0 | 54/186(29.0) | 154/511 (30.1) | 1.1 (-6.9; 9.1) | 0.851 |
| Unprotected sex (past 12 months): No | 129/237 (54.4) | 85 / 354(24.0) | -30.4 ( -38.5; -22.3) | 0 | 106/236(44.9) | 124/359 (34.5) | -10.4 (-18.8; -2) | 0.014 |
| Unprotected sex (past 12 months): Yes | 66/184 (35.9) | 109/510(21.4) | -14.5 ( -22.7; -6.3) | 0 | 54/186(29.0) | 154/511 (30.1) | 1.1 (-6.9; 9.1) | 0.851 |
| Received money, goods, or services for sex: Yes | 24/58 (41.4) | 175/774(22.6) | -18.8 ( -32.7; -4.8) | 0.002 | 21/59(35.6) | 234/777(30.1) | -5.5 ( -19; 8.1) | 0.463 |
| Received money, goods, or services for sex: No | 171/363 (47.1) | 21/117(17.9) | -29.2 ( -38.4; -19.9) | 0 | 138/362(38.1) | 55/120(45.8) | 7.7 ( -3.1; 18.5) | 0.166 |
| **Health service use & risk perception** |  |  |  |  |  |  |  |  |
| Self-reported Sexual transmitted infections (STI): Yes | 42/101 (41.6) | 118/476 (24.8) | -16.8 (-27.8; -5.8) | 0.001 | 23/99(23.2) | 123/482 (25.5) | 2.3 (-7.5; 12.1) | 0.726 |
| Self-reported STI: No | 153/321 (47.7) | 80/440 (18.2) | -29.5 (-36.3; -22.7) | 0 | 137/323(42.4) | 175/441 (39.70) | -2.7 (-10.1; 4.6) | 0.494 |
| HIV test in the last month: Yes | 53/99 (53.5) | 86/580 (14.8) | -38.7 (-49.5; -27.9) | 0 | 43/97(44.3) | 212/585 (36.2) | -8.1 (-19.3; 3.1) | 0.158 |
| HIV test in the last month: No | 141/320 (44.1) | 109/314 (34.7) | -9.3 (-17.2; -1.5) | 0.02 | 117/322(36.3) | 81/316 (25.6) | -10.7 (-18.1; -3.3) | 0.005 |
| HIV risk perception: No/low risk | 14/84 (16.7) | 4/114 (3.5) | -13.2 (-22.8; -3.5) | 0.003 | 18/91(19.8) | 49/116 (42.2) | 22.5 (9.3; 35.6) | 0.001 |
| HIV risk perception: Moderate/high risk | 41/146 (28.1) | 37/543(6.8) | -21.3 (-29.3; -13.2) | 0 | 40/152 (26.3) | 161/546 (29.5) | 3.2 ( -5.2; 11.6) | 0.508 |
| HIV risk perception: Refused | 16/64 (25.0) | 14/103(13.6) | -11.4 (-25.2; 2.4) | 0.097 | 11/64 (17.2) | 24/103 (23.3) | 6.1 ( -7.5; 19.7) | 0.454 |
| Sought healthcare (past 12 months): Yes | 84/159 (52.8) | 126/505(25.0) | -27.9 (-36.9; -18.8) | 0 | 56/158 (35.4) | 167/510 (32.7) | -2.7 ( -11.6; 6.2) | 0.595 |
| Sought healthcare (past 12 months): No | 111/263 (42.8) | 72/411(17.5) | -24.7 (-32; -17.4) | 0 | 104/264 (39.4) | 131/413 (31.7) | -7.7 ( -15.4; 0) | 0.05 |

<sup>a</sup> Denominators vary across variables due to missing data and survey eligibility criteria.
<sup>b</sup> p-values derived from Chi-square tests for equality of proportions between 2014 and 2023.
<sup>c</sup> All estimates represent unweighted sample proportions based on raw counts; RDS weighting was not applied.
<sup>d</sup> Categories with small cell counts ( $n < 20$ ) should be interpreted with caution.

#### HIV prevalence

prevalence among PWID in Mozambique between 2014 and 2023. Contrary to the initial expectation of increased HIV burden, the unadjusted descriptive results showed significant reductions across almost all demographic, behavioural and service-use subgroups (Table 1).

Among PWID participants aged ≥25 years, HIV prevalence declined markedly from 55.7% in 2014 to 26.3% in 2023 (p < 0.001). A non-significant increase was observed among younger PWID aged 16–24 years (6.2% to 13.0%; p = 0.126). A similar pattern of reduction was evident across sex categories: prevalence decreased significantly among men (45.7% to 16.7%; p < 0.001) and declined non-significantly among women (57.9% to 43.2%; p = 0.257).

Declines were also observed across education levels. HIV prevalence fell from 52.0% to 30.6% among PWID with no formal or primary education (p < 0.001) and from 42.1% to 17.6% among those with secondary or higher education (p < 0.001). Across marital status, prevalence dropped from 44.9% to 18.0% among those never married (p < 0.001), remained stable among those married or living in union (39.5% to 34.0%; p = 0.477), and declined significantly among those in other marital categories (56.2% to 28.0%; p < 0.001). Geographically, Maputo City recorded one of the largest reductions, with HIV prevalence decreasing from 56.6% to 28.0% (p < 0.001). In Nampula/Nacala, a smaller, non-significant decline was observed (20.0% to 14.2%; p = 0.158).

Declines in HIV prevalence was also evident across injecting-related characteristics. Among daily injectors, prevalence dropped sharply from 58.0% to 21.3% (p < 0.001). Substantial reductions were also observed among participants reporting recent syringe sharing (75.0% to 21.8%; p < 0.001) and among those who reported never sharing syringes (43.7% to 20.7%; p < 0.001).

Regarding sexual behaviors, HIV prevalence declined significantly across all categories of sexual partners. Among PWID with 0–1 partners, prevalence fell from 50.6% to 24.4% (p < 0.001); among those with two partners, from 46.4% to 17.1% (p < 0.001); and among those with three or more partners, from 42.3% to 21.0% (p < 0.001). Prevalence also decreased significantly among participants reporting unprotected sex (35.9% to 21.4%; p < 0.001) and those reporting no unprotected sex (54.4% to 24.0%; p < 0.001).

#### HCV prevalence

HCV prevalence showed heterogeneous patterns across demographic and behavioural groups (Table 1). Among younger PWID aged 16–24 years, prevalence increased sharply from 7.3% to 38.7% (p < 0.001), while a significant decline was observed among those aged ≥25 years, from 45.3% to 28.8% (p < 0.001). Prevalence also decreased among women (43.8% to 16.4%; p = 0.019) and among PWID with no formal/primary education (43.0% to 27.8%; p = 0.001), whereas no change occurred among those with secondary or higher education (p = 0.968).

Geographic trends diverged strongly: Maputo City recorded a substantial reduction in HCV prevalence (49.3% to 18.7%; p < 0.001), whereas Nampula/Nacala experienced a significant increase (11.7% to 48.1%; p < 0.001).

Injecting-related behaviors also reflect mixed patterns. Prevalence declined significantly among daily injectors (51.8% to 32.6%; p < 0.001), among those reporting no access to new syringes (40.0% to 18.4%; p = 0.005), and among those who did not share syringes (36.3% to 14.9%; p < 0.001). Among participants who shared syringes, HCV prevalence decreased from 75.0% to 38.9% (p = 0.003).

Sexual behavior patterns showed minimal change. Prevalence remained stable among individuals with 0–1 partners (p = 0.917) and two partners (p = 0.412) but declined slightly among those with ≥3 partners (36.4% to 27.2%; p = 0.026). No significant differences were found for transactional sex.

### Changes in Prevalence by City

Table 2 presents change in the odds of HIV and HCV infection between 2014 and 2023 for Maputo and Nampula. For HIV, a marked reduction was observed in Maputo, where the odds of infection in 2023 were 70% lower than in 2014 (COR = 0.3; 95% CI: 0.2–0.4; p<0.001). In Nampula, the reduction was not statistically significant (COR = 0.6; 95% CI: 0.4–1.1; p = 0.122). When both cities were combined, the overall odds of HIV infection also decreased significantly (COR = 0.3; 95% CI: 0.3–0.4; p < 0.001).

**Table 2.**
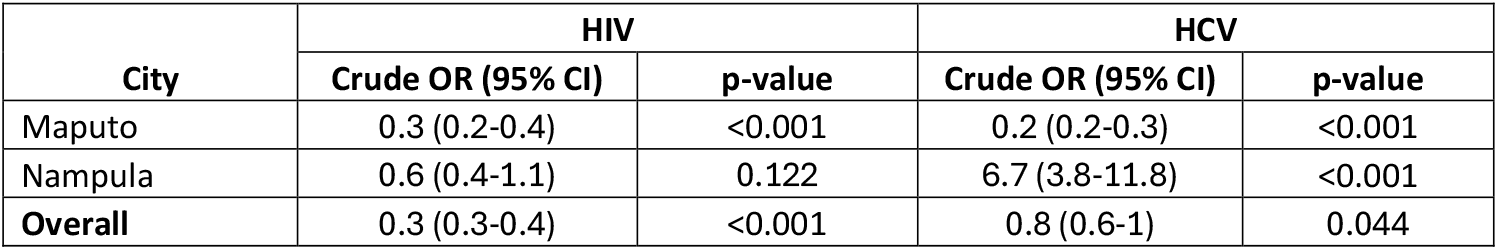
Changes in HIV and HCV prevalence among PWID by City.

| City | HIV |  | HCV |  |
| --- | --- | --- | --- | --- |
|  | Crude OR (95% CI) | p-value | Crude OR (95% CI) | p-value |
| Maputo | 0.3 (0.2-0.4) | <0.001 | 0.2 (0.2-0.3) | <0.001 |
| Nampula | 0.6 (0.4-1.1) | 0.122 | 6.7 (3.8-11.8) | <0.001 |
| **Overall** | 0.3 (0.3-0.4) | <0.001 | 0.8 (0.6-1) | 0.044 |

For HCV, trends differed sharply by city. In Maputo, the odds of HCV infection declined by approximately 80% (COR = 0.2; 95% CI: 0.2–0.3; p < 0.001). In contrast, Nampula experienced a substantial increase, with PWID nearly seven times more likely to be infected in 2023 compared with 2014 (COR = 6.7; 95% CI: 3.8–11.8; p < 0.001). When both cities were combined, the pooled estimate suggested a modest reduction overall, although this was only marginally significant (COR = 0.8; 95% CI: 0.6–1.0; p = 0.044).

### Multivariable logistic regression analysis identified several factors significantly associated with HIV and HCV infections among PWID in Mozambique

#### Risk factors for HIV

Table 3 shows that age, sex, marital status, injection behaviour and HIV testing history were independently associated with HIV infection. PWID aged ≥25 years had significantly higher odds of HIV infection compared with younger PWID (AOR = 12.2; 95% CI: 4.6–32.5; p < 0.001). A significant interaction with survey year indicated that this age effect decreased substantially in 2023 (interaction AOR = 0.2; 95% CI: 0.1–0.7; p = 0.009).

**Table 3.**
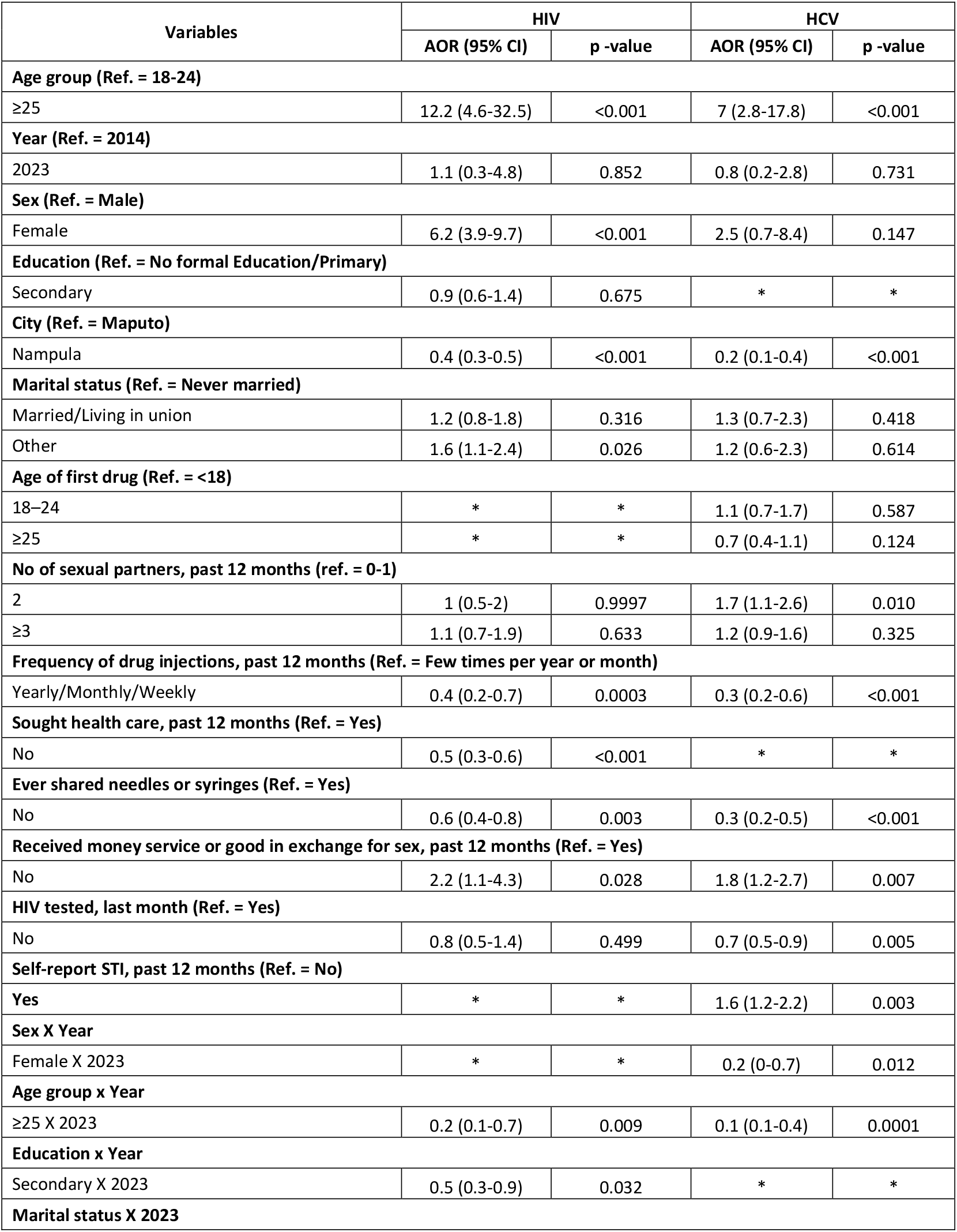
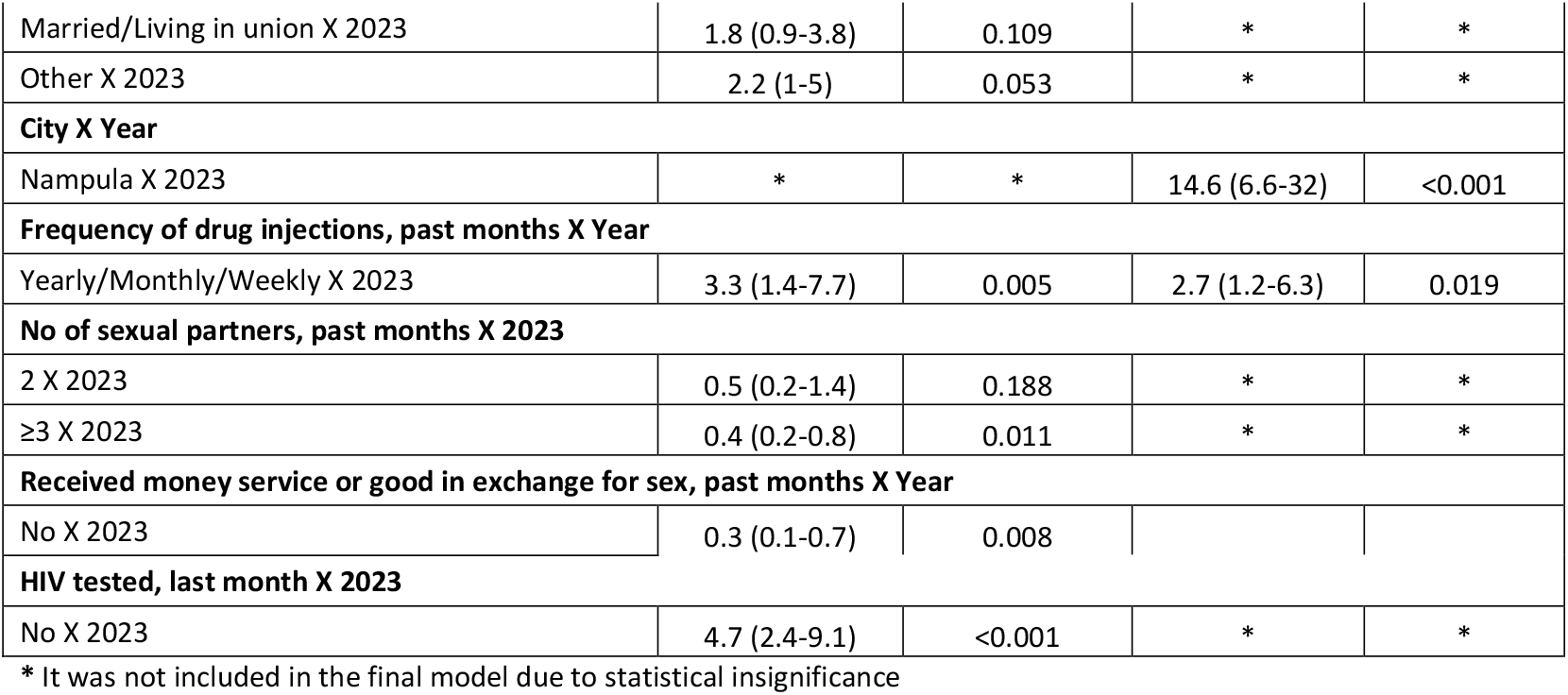
Multivariate logistic regression parameter estimates. The estimates are presented in the odds ratio (OR) scale accompanied by 95 % confidence interval in brackets.

| Variables | HIV |  | HCV |  |
| --- | --- | --- | --- | --- |
|  | AOR (95% CI) | p -value | AOR (95% CI) | p -value |
| **Age group (Ref. = 18-24)** |  |  |  |  |
| ≥25 | 12.2 (4.6-32.5) | <0.001 | 7 (2.8-17.8) | <0.001 |
| **Year (Ref. = 2014)** |  |  |  |  |
| 2023 | 1.1 (0.3-4.8) | 0.852 | 0.8 (0.2-2.8) | 0.731 |
| **Sex (Ref. = Male)** |  |  |  |  |
| Female | 6.2 (3.9-9.7) | <0.001 | 2.5 (0.7-8.4) | 0.147 |
| **Education (Ref. = No formal Education/Primary)** |  |  |  |  |
| Secondary | 0.9 (0.6-1.4) | 0.675 | * | * |
| **City (Ref. = Maputo)** |  |  |  |  |
| Nampula | 0.4 (0.3-0.5) | <0.001 | 0.2 (0.1-0.4) | <0.001 |
| **Marital status (Ref. = Never married)** |  |  |  |  |
| Married/Living in union | 1.2 (0.8-1.8) | 0.316 | 1.3 (0.7-2.3) | 0.418 |
| Other | 1.6 (1.1-2.4) | 0.026 | 1.2 (0.6-2.3) | 0.614 |
| **Age of first drug (Ref. = <18)** |  |  |  |  |
| 18–24 | * | * | 1.1 (0.7-1.7) | 0.587 |
| ≥25 | * | * | 0.7 (0.4-1.1) | 0.124 |
| **No of sexual partners, past 12 months (ref. = 0-1)** |  |  |  |  |
| 2 | 1 (0.5-2) | 0.9997 | 1.7 (1.1-2.6) | 0.010 |
| ≥3 | 1.1 (0.7-1.9) | 0.633 | 1.2 (0.9-1.6) | 0.325 |
| **Frequency of drug injections, past 12 months (Ref. = Few times per year or month)** |  |  |  |  |
| Yearly/Monthly/Weekly | 0.4 (0.2-0.7) | 0.0003 | 0.3 (0.2-0.6) | <0.001 |
| **Sought health care, past 12 months (Ref. = Yes)** |  |  |  |  |
| No | 0.5 (0.3-0.6) | <0.001 | * | * |
| **Ever shared needles or syringes (Ref. = Yes)** |  |  |  |  |
| No | 0.6 (0.4-0.8) | 0.003 | 0.3 (0.2-0.5) | <0.001 |
| **Received money service or good in exchange for sex, past 12 months (Ref. = Yes)** |  |  |  |  |
| No | 2.2 (1.1-4.3) | 0.028 | 1.8 (1.2-2.7) | 0.007 |
| **HIV tested, last month (Ref. = Yes)** |  |  |  |  |
| No | 0.8 (0.5-1.4) | 0.499 | 0.7 (0.5-0.9) | 0.005 |
| **Self-report STI, past 12 months (Ref. = No)** |  |  |  |  |
| Yes | * | * | 1.6 (1.2-2.2) | 0.003 |
| **Sex X Year** |  |  |  |  |
| Female X 2023 | * | * | 0.2 (0-0.7) | 0.012 |
| **Age group x Year** |  |  |  |  |
| ≥25 X 2023 | 0.2 (0.1-0.7) | 0.009 | 0.1 (0.1-0.4) | 0.0001 |
| **Education x Year** |  |  |  |  |
| Secondary X 2023 | 0.5 (0.3-0.9) | 0.032 | * | * |
| **Marital status X 2023** |  |  |  |  |
| Married/Living in union X 2023 | 1.8 (0.9-3.8) | 0.109 | * | * |
| Other X 2023 | 2.2 (1-5) | 0.053 | * | * |
| **City X Year** |  |  |  |  |
| Nampula X 2023 | * | * | 14.6 (6.6-32) | <0.001 |
| **Frequency of drug injections, past months X Year** |  |  |  |  |
| Yearly/Monthly/Weekly X 2023 | 3.3 (1.4-7.7) | 0.005 | 2.7 (1.2-6.3) | 0.019 |
| **No of sexual partners, past months X 2023** |  |  |  |  |
| 2 X 2023 | 0.5 (0.2-1.4) | 0.188 | * | * |
| ≥3 X 2023 | 0.4 (0.2-0.8) | 0.011 | * | * |
| **Received money service or good in exchange for sex, past months X Year** |  |  |  |  |
| No X 2023 | 0.3 (0.1-0.7) | 0.008 |  |  |
| **HIV tested, last month X 2023** |  |  |  |  |
| No X 2023 | 4.7 (2.4-9.1) | <0.001 | * | * |

\* It was not included in the final model due to statistical insignificance

Female PWID had consistently higher odds of HIV infection (AOR = 6.2; 95% CI: 3.9–9.7; p < 0.001). Marital status also showed an association: participants in the “other” category (widowed/divorced/separated) had higher odds of HIV infection compared with those never married (AOR = 1.6; 95% CI: 1.1–2.4; p = 0.026).

Geographically, PWID residing in Nampula had significantly lower odds of HIV infection than those in Maputo (AOR = 0.4; 95% CI: 0.3–0.5; p < 0.001). Lower injection frequency (weekly/monthly/yearly vs. few times per month/year) was protective (AOR = 0.4; 95% CI: 0.2–0.7; p = 0.0003), as was not sharing needles or syringes (AOR = 0.6; 95% CI: 0.4–0.8; p = 0.003).

Participants who did not report transactional sex had higher odds of HIV infection (AOR = 2.2; 95% CI: 1.1–4.3; p = 0.028), and this effect differed significantly in 2023 (interaction AOR = 0.3; 95% CI: 0.1– 0.7; p = 0.008).

Not having tested for HIV in the last month was not associated with HIV infection in the pooled model (AOR = 0.8; p = 0.499), but the interaction term showed that in 2023 this group had higher odds of HIV infection (AOR = 4.7; 95% CI: 2.4–9.1; p < 0.001).

#### Risk factors for HCV

For HCV, older age was strongly associated with infection (AOR = 7.0; 95% CI: 2.8–17.8; p < 0.001). Residing in Nampula was protective overall (AOR = 0.2; 95% CI: 0.1–0.4; p < 0.001), although the interaction term indicated a dramatic increase in 2023 (AOR = 14.6; 95% CI: 6.6–32.0; p < 0.001).

Having two sexual partners in the past 12 months was associated with higher odds of HCV infection (AOR = 1.7; 95% CI: 1.1–2.6; p = 0.010). Safer injecting practices were protective: PWID who did not share needles had significantly lower odds of HCV infection (AOR = 0.3; 95% CI: 0.2–0.5; p < 0.001), and lower injection frequency was also protective (AOR = 0.3; p < 0.001), with a significant interaction indicating variation in 2023 (AOR = 2.7; p = 0.019).

Not engaging in transactional sex was associated with increased odds of HCV (AOR = 1.8; 95% CI: 1.2– 2.7; p = 0.007). Participants reporting STI in the past year had higher odds of HCV infection (AOR = 1.6; 95% CI: 1.2–2.2; p = 0.003).

Significant interactions also indicated reduced odds among women in 2023 (AOR = 0.2; 95% CI: 0.0– 0.7; p = 0.012) and among older PWID in 2023 (AOR = 0.1; 95% CI: 0.1–0.4; p = 0.0001).

## Discussion

This study provides the first comparative assessment of HIV and HCV trends among PWID in Mozambique across a 10-year interval, revealing strong declines in HIV prevalence but worrying and divergent patterns in HCV infection, particularly in Nampula. These findings highlight meaningful progress in some areas of harm reduction, while also signaling emerging vulnerabilities requiring urgent attention

HIV prevalence declined across almost all demographic and behavioural groups, with Maputo showing a marked reduction over time. In contrast, prevalence in Nampula remained stable, with no statistically significant change, highlighting geographic differences in epidemic dynamics and service coverage. The marked reductions among daily injectors, individuals who reported sharing syringes, and those with multiple sexual partners suggest structural improvements in access to sterile injecting equipment, HIV testing, and ART uptake [11,12]. Similar patterns have been documented in settings where partial harm reduction coverage, such as expanded HIV testing or improved linkage to ART, reduced HIV transmission even in the absence of fully comprehensive services [13,14]. In multivariable analyses, older PWID (≥25 years) had substantially higher odds of HIV infection, but this age effect attenuated over time, indicating a partial shift of HIV risk towards younger PWID in 2023.

Nevertheless, HIV risk remains unevenly distributed. Female PWID continued to experience disproportionately higher odds of infection, reflecting gendered vulnerabilities linked to stigma, violence, economic dependence and reduced access to health services. These findings align with evidence from sub-Saharan Africa showing that women who inject drugs experience higher HIV burden due to overlapping structural risks such as gender based violence and limited access to health care service [12,15,16]. Across both survey years, PWID in Nampula had consistently lower odds of HIV infection than those in Maputo, suggesting site-specific differences in epidemic maturity, service availability or injecting networks that warrant further investigation. In addition, PWID who had not tested for HIV in the last month had markedly higher odds of HIV infection in 2023, underscoring persistent gaps in regular testing and missed opportunities for timely diagnosis and linkage to care.

In contrast to HIV, HCV trends revealed strong geographic polarization. Maputo experienced a substantial reduction in HCV prevalence, whereas Nampula recorded a dramatic increase, further confirmed by the very large interaction effect in the regression model (AOR 14.6). Such rapid rises are consistent with HCV outbreaks in emerging or expanding injecting networks, where insufficient coverage of sterile injecting equipment can accelerate transmission [14,17]. The sharp increase in HCV prevalence among younger PWID (18–24 years), together with the strong overall association between older age and HCV in the pooled model, points to a changing age profile of infection, with new cohorts of younger injectors entering high-risk networks. The sharp increase among younger PWID also suggests recent entry into injecting practices and lower awareness or access to safer injecting supplies[18].

The divergence between HIV and HCV trajectories, declining HIV alongside rising HCV in certain subgroups, reflects the biological and programmatic reality that HCV is far more efficiently transmitted via injecting equipment and requires higher coverage of sterile supplies to interrupt transmission [19,20]. This pattern implies that existing services may be adequate to suppress HIV but insufficient to control HCV in regions such as Nampula.

Several protective factors identified in the multivariable models reinforce the need to strengthen harm reduction. Lower injection frequency and not sharing needles significantly reduced odds of both HIV and HCV infection, while accessing healthcare was protective for HIV. The associations linking absence of transactional sex to higher odds of both infections likely reflect underreporting or complex social risk structures rather than causal mechanisms, warranting further qualitative research [12,21,22].

This study has several limitations that should be considered when interpreting the findings. First, both BBS rounds used cross-sectional designs, which preclude causal inference and do not allow individual-level follow-up; observed differences over time may partly reflect changes in the composition of PWID rather than true cohort effects. Second, although recruitment used RDS, we applied unweighted analyses because data were aggregated across cities and RDS estimators are only valid within single recruitment networks; this may introduce selection bias and limit the generalizability of prevalence estimates. Third, behavioral data, including sexual practices, injecting behaviors and transactional sex, were self-reported and therefore subject to recall and social desirability bias, which could lead to misclassification of key exposures. Fourth, HCV status was based on rapid antibody tests and did not distinguish between past resolved infection and current viremic infection, potentially overestimating active HCV burden. Finally, some subgroups had small cell sizes, particularly women and specific behavioral categories, resulting in wide confidence intervals and reduced statistical power; these estimates should be interpreted with caution. Despite these limitations, the study provides longitudinally comparable evidence on HIV and HCV trends among PWID in Mozambique and offers critical insights to guide harm reduction policy and programming.

## Conclusion

This study provides the first longitudinally comparable evidence on HIV and HCV epidemiological trends among PWID in Mozambique, revealing substantial yet uneven progress over the past decade. HIV prevalence declined sharply across most demographic and behavioral groups, particularly in Maputo, and notable reductions were observed even among traditionally high-risk subgroups such as daily injectors and individuals who reported syringe sharing. These patterns suggest improvements in access to sterile injecting equipment, HIV testing and ART uptake. However, persistent gender disparities, the higher burden among women and gaps in regular HIV testing highlight ongoing structural vulnerabilities that require targeted, gender-responsive interventions.

In contrast, HCV trends expose widening geographic and generational risks. The marked increase in HCV prevalence in Nampula, alongside evidence of a younger cohort entering high-risk injecting networks, indicates an emerging and potentially expanding HCV transmission hotspot. The divergence between HIV and HCV trajectories highlights that current services, while effective for HIV control, are insufficient to prevent HCV transmission.

Taken together, the findings call for differentiated, city-specific harm reduction strategies, with expansion of HCV testing and linkage to care in Nampula, continued investment in prevention and ART integration in Maputo, and strengthened behavioral, structural and gender-responsive interventions across all sites. Scaling up comprehensive harm reduction is essential to prevent further HCV transmission and to consolidate gains achieved in HIV control. Sustained investment in these evidence-based interventions will be critical for Mozambique to advance towards global HIV and viral hepatitis elimination goals and to ensure that PWID are no longer left behind.

## Data Availability

All study information and datasets are fully accessible at the Mozambique National Institute of Health (INS) data repository for researchers who meet the criteria for accessing confidential data. The data originate from the BBS studies, and the authors can be contacted through: https://ins.gov.mz/institucional/unidade-organicas/direccoes/direccao-de-inqueritos-e-observacao-de-saude/bases-de-dados/

## Acknowledgements

The authors extend their gratitude to all individuals who contributed to the planning, execution, analysis, and dissemination of the BBS results, including investigators, study teams, Technical Working Groups, Non-Governmental Organizations, members of civil society, and the National HIV/STD program.

## Authors’ contributions

**Conceptualization:** Áuria Ribeiro Banze.

**Data curation:** Áuria Ribeiro Banze, Rachid Muleia, Diogo Chavana, Samuel Nuvunga, Cynthia Semá Baltazar.

**Formal analysis:** Rachid Muleia.

**Funding acquisitio**n: Áuria Ribeiro Banze, Cynthia Semá Baltazar.

**Investigation:** Áuria Ribeiro Banze, Cynthia Semá Baltazar

**Methodology:** Áuria Ribeiro Banze, Rachid Muleia, Cynthia Semá Baltazar.

**Project administration:** Áuria Ribeiro Banze, Lúcia Muioche, Cynthia Semá Baltazar.

**Resources:** Áuria Ribeiro Banze, Cynthia Semá Baltazar

**Software**: Diogo Chavana

**Supervision:** Lúcia Muioche, Samuel Nuvunga, Gércio Cuamba, Manuel Condula, Stélio Craveirinha, Diogo Chavana, Ana Mutola Jemuce, Valdo Mega, Daniel Chilaúle, Maria Simbine, Carlos Botão, Nália Ismael

**Validation:** Áuria Ribeiro Banze, Rachid Muleia and Cynthia Semá Baltazar.

**Visualization:** Diogo Chavana, Rachid Muleia, Samuel Nuvunga.

**Writing– original draft:** Áuria Ribeiro Banze, Rachid Muleia, Lúcia Muioche, Cynthia Semá Baltazar.

**Writing– review & editing:** Áuria Ribeiro Banze, Rachid Muleia, Lúcia Muioche, Samuel Nuvunga, Gércio Cuamba, Manuel Condula, Stélio Craveirinha, Diogo Chavana, Ana Mutola Jemuce, Valdo Mega, Daniel Chilaúle, Maria Simbine, Carlos Botão, Nália Ismael, Cynthia Semá Baltazar.

## References

1. WHO. Data on the size of the HIV epidemic [Internet]. [cited 2025 Nov 16]. https://www.who.int/data/gho/data/themes/hiv-aids/data-on-the-size-of-the-hiv-aids-epidemic?utm_source=chatgpt.com. Accessed 16 Nov 2025

2. Hepatitis C [Internet]. [cited 2025 Nov 16]. https://www.who.int/news-room/fact-sheets/detail/hepatitis-c. Accessed 16 Nov 2025

3. Semá Baltazar C, Horth R, Boothe M, Sathane I, Young P, Chitsondzo Langa D, et al. High prevalence of HIV, HBsAg and anti-HCV positivity among people who injected drugs: results of the first bio-behavioral survey using respondent-driven sampling in two urban areas in Mozambique. BMC Infect Dis. 2019;19:1022. 10.1186/s12879-019-4655-2

4. World Drug Report 2025 - Special Points of Interest [Internet]. United Nations : Office on Drugs and Crime. [cited 2025 Sep 7]. http://www.unodc.org/unodc/en/data-and-analysis/world-drug-report-2025-special-points-of-interest.html. Accessed 7 Sep 2025

5. UNAIDS. HIV AND PEOPLE WHO USE DRUGS - Fact sheet [Internet]. 2024 p. 7. https://www.unaids.org/sites/default/files/media_asset/02-hiv-human-rights-factsheet-people-who-use-drugs_en.pdf?utm_source=chatgpt.com. Accessed 18 Aug 2025

6. WHO. Global HIV, Hepatitis and STIs Programmes [Internet]. World Health Organization; https://www.who.int/teams/global-hiv-hepatitis-and-stis-programmes/populations/people-who-inject-drugs. Accessed 16 Aug 2025

7. INS. Inquérito Nacional sobre o impacto do HIV e SIDA em Moçambique. INSIDA 2021 [Internet]. 2022. https://www.ins.gov.mz/wp-content/uploads/2022/12/53059_14_INSIDA_Summary-sheet_POR.pdf. Accessed 18 Aug 2025

8. CNCS. Plano Estratégico Nacional de Resposta ao HIV e SIDA – PEN V (2021 - 2025) [Internet]. Conselho Nacional de Combate ao HIV/SIDA; 2021 p. 100. https://www.unaids.org/en/resources/documents/2023/2022_unaids_data

9. Instituto Nacional de Saúde. Inquérito Biológico e Comportamental entre Pessoas que Injectam Drogas (BBS-PID) – 2023 [Internet]. Instituto Nacional de Saúde. [cited 2025 Oct 26]. https://ins.gov.mz/relatorios-cientificos/. Accessed 26 Oct 2025

10. Semá Baltazar C, Boothe M, Kellogg T, Ricardo P, Sathane I, Fazito E, et al. Prevalence and risk factors associated with HIV/hepatitis B and HIV/hepatitis C co-infections among people who inject drugs in Mozambique. BMC Public Health. 2020;20:851. 10.1186/s12889-020-09012-w

11. UNAIDS. Global HIV & AIDS statistics — Fact sheet | UNAIDS [Internet]. [cited 2025 Oct 26]. https://www.unaids.org/en/resources/fact-sheet. Accessed 26 Oct 2025

12. Banze ÁR, Botão C, Muamine E, Condula, Manuel. Understanding HIV vulnerability among women who inject drugs in Mozambique, 2023 | Harm Reduction Journal | Full Text [Internet]. [cited 2025 Oct 26]. https://harmreductionjournal.biomedcentral.com/articles/10.1186/s12954-025-01204-0?utm_source=chatgpt.com. Accessed 26 Oct 2025

13. Nelson PK, Mathers BM, Cowie B, Hagan H, Des Jarlais D, Horyniak D, et al. Global epidemiology of hepatitis B and hepatitis C in people who inject drugs: results of systematic reviews. Lancet. 2011;378:571–83. 10.1016/S0140-6736(11)61097-0

14. Degenhardt L, Peacock A, Colledge S, Leung J, Grebely J, Vickerman P, et al. Global prevalence of injecting drug use and sociodemographic characteristics and prevalence of HIV, HBV, and HCV in people who inject drugs: a multistage systematic review. Lancet Glob Health. 2017;5:e1192–207. 10.1016/S2214-109X(17)30375-3

15. Otanga H, Jeneby F, Handulle M, Busz M, Sumnall H, Van Hout MC. Gender based violence against women who use drugs (WWUD) in Kenya: Experiences and Policy Directions. Kenya Policy Briefs. 2022;

16. Aung SWKH, Kingston H, Mbogo LW, Sambai B, Monroe-Wise A, Ludwig-Barron NT, et al. Prevalence and correlates of violence among sexual and injecting partners of people who inject drugs living with HIV in Kenya: a cross-sectional study. Harm Reduct J. 2023;20:164. 10.1186/s12954-023-00895-7

17. Kassa GM, Lim AG, Tamiru MT, Alamneh TS, Vickerman P, Dagne E, et al. Risk Factors for Hepatitis C Virus Among the General Population in Sub-Saharan Africa—An Analysis of Systematic Review Data. J Viral Hepat. 2025;32:e70065. 10.1111/jvh.70065

18. Impact of the US funding cuts on harm reduction - Harm Reduction International [Internet]. [cited 2025 Oct 26]. https://hri.global/publications/impact-of-the-us-funding-cuts-on-harm-reduction/. Accessed 26 Oct 2025

19. Platt L, Minozzi S, Reed J, Vickerman P, Hagan H, French C, et al. Needle and syringe programmes and opioid substitution therapy for preventing HCV transmission among people who inject drugs: findings from a Cochrane Review and meta-analysis. Addiction. 2018;113:545–63. 10.1111/add.14012

20. Fernandes RM, Cary M, Duarte G, Jesus G, Alarcão J, Torre C, et al. Effectiveness of needle and syringe Programmes in people who inject drugs – An overview of systematic reviews. BMC Public Health. 2017;17:309. 10.1186/s12889-017-4210-2

21. Unveiling triple vulnerability among Mozambican female sex workers—Stigma, physical violence and sexual violence | PLOS One [Internet]. [cited 2025 Oct 26]. https://journals.plos.org/plosone/article?id=10.1371/journal.pone.0312550. Accessed 26 Oct 2025

22. Situational assessment and epidemiology of HIV, HBV and HCV among people who use and inject drugs in Ghana | PLOS One [Internet]. [cited 2025 Oct 26]. https://journals.plos.org/plosone/article?id=10.1371/journal.pone.0305923. Accessed 26 Oct 2025

